# Global Research Trends, Hotspots, Impacts, and Emerging Developments in Intelligent Operating Room Nursing: A 10-Year Bibliometric Analysis

**DOI:** 10.1101/2025.06.18.25329778

**Authors:** Yang Yu, Zhili Rao, Wen Zheng, Ting Su, Xin Wen, Lijun Yao, Weixi Zheng, Ying Wang, Yaling Wang

## Abstract

**Background and Objectives:** The integration of intelligent technologies in operating room nursing represents a rapidly evolving field requiring systematic analysis to understand global research trends and development patterns. This study aimed to comprehensively analyze worldwide research trends, identify key contributors, and reveal emerging developments in intelligent operating room nursing through bibliometric analysis.

**Methods:** A comprehensive literature search was conducted across six international databases (Web of Science, Scopus, PubMed, Embase, Cochrane Library, and CINAHL) from January 2015 to April 2025. Following rigorous screening criteria, 291 eligible articles were analyzed using R software with bibliometrix package, VOSviewer, and CiteSpace for bibliometric analysis, network visualization, and thematic evolution assessment.

**Results:** Publication output increased from 16 articles in 2015 to 49 in 2024, with 45.4% of total publications concentrated in 2022-2024. The United States dominated with 132 publications, followed by Germany and the United Kingdom (33 each). Technical University of Munich led institutional contributions with 7 publications. Six major research themes emerged: operating room management, robotic technology, nursing practice, patient safety, quality improvement, and intelligent algorithms. Keyword analysis revealed evolution from early focus on operating room scheduling and multi-objective optimization (2017) to deep learning applications (2020-2023), natural language processing (2023), and robotic scrub nurse development (2023-2025).

**Conclusions:** The field demonstrates rapid growth with clear thematic evolution toward human-machine collaboration systems. Future research should prioritize comparative effectiveness studies, implementation science methodologies, and global health equity considerations to ensure widespread clinical translation.

## 1. Introduction

Over the course of several decades, intelligent healthcare systems have evolved to incorporate increasingly sophisticated technologies that enhance human capabilities in clinical settings[1–3]. Similar to other healthcare domains, intelligent operating room nursing encompasses numerous specialized areas, including smart monitoring systems, predictive analytics, and automated documentation[4]. The utilization of real-time data to identify patterns that can be applied to evaluate patient status is known as intelligent monitoring[5, 6]. Instead of relying on manual observation alone, these intelligent tools can be actively integrated into perioperative nursing workflows to personalize patient care during surgery[7–9]. A system’s ability to capture and process information from various operating room sensors and equipment is known as integrated data management[6], creating new possibilities for patient monitoring, procedural efficiency, and safety enhancement[9, 10]. Technology-driven innovations are increasingly implemented in surgical workflow management, intraoperative decision support, and resource optimization, enabling more responsive care coordination, improved surgical outcomes, and reduced procedural complications, advancing perioperative practice toward data-informed interventions, enhanced team communication, and optimized patient safety protocols[11]. As intelligent technologies continue to reshape surgical environments, understanding the evolution of smart systems research in operating room nursing is essential for identifying development trajectories, implementation challenges, and pioneering institutions in this emerging field[12, 13].

Bibliometric analysis provides a powerful methodological framework for comprehensively examining global research developments, detecting emerging areas, and identifying influential studies across various surgical specialties[14–17]. Previous bibliometric investigations have explored digital technologies or smart systems in general nursing or surgical contexts, or institution-specific implementations, but the global and thorough assessments of intelligent technology integration in operating room nursing remain scarce[18–20]. This study presents a comprehensive worldwide analysis of intelligent technology research trends in operating room nursing from 2015 to 2025. This study aims to (1) evaluate annual publication output and citation patterns, (2) identify key contributors including prominent researchers, countries, journals, and institutions, and (3) reveal current research frontiers in the application of intelligent technologies in operating room nursing by examining author keywords and research themes.

Through a comprehensive and data-driven analysis of intelligent operating room literature, this study provides valuable insights for clinical nursing researchers, surgical teams, hospital management, and medical device manufacturers. Understanding these evolving research patterns can guide future technological developments, ensuring that smart operating room innovations continue to enhance surgical outcomes, improve patient care, and optimize perioperative workflows.

## 2. METHODS

### 2.1 Data sources and search strategy

We conducted a comprehensive literature search across multiple international databases to ensure comprehensive coverage of relevant publications. The English literature search was performed on the Web of Science Core Collection (WoSCC) database (https://www.webofscience.com/wos/woscc/basic-search), Scopus database (http://www.scopus.com), PubMed database (https://pubmed.ncbi.nlm.nih.gov/), Embase database (https://www.embase.com/), Cochrane Library (https://www.cochranelibrary.com/), and CINAHL Complete database (https://www.ebscohost.com/nursing/products/cinahl-databases).

The search strategy employed was “(TS = (smart operating room) OR TS = (intelligent operating room) OR TS = (operating room intelligence) OR TS = (OR automation) OR TS = (surgical robotics) OR TS = (operating theatre intelligence)) AND (TS = (artificial intelligence) OR TS = (machine learning) OR TS = (deep learning) OR TS = (automation) OR TS = (digital surgery))” for relevant publications. The reference types included “article” and “review” publications. The temporal scope encompassed publications from “January 1, 2015–April 31, 2025”. All data were acquired on May 1, 2025, to minimize bias caused by database updates.

A comprehensive search of multiple databases yielded a total of 923 publications. The database sources included Web of Science (179 articles), CINAHL (65 articles), COCHRANE (136 articles), EMBASE (78 articles), PubMed (157 articles), and Scopus (308 articles). Following the initial search, a rigorous screening process was conducted using specific exclusion criteria (**Figure 1**). A total of 632 articles were excluded: 386 duplicates, 17 non-English articles, 11 withdrawn publications, and 219 articles unrelated to the research topic. The remaining 291 eligible articles were imported into EndNote for data cleaning and further analysis. Subsequently, bibliometric and visualization analyses were performed across multiple dimensions, including publications, citations, countries, institutions, journals, and authors. This systematic approach ensured a comprehensive evaluation of the current research landscape in intelligent operating room systems[21].

**Figure 1.**
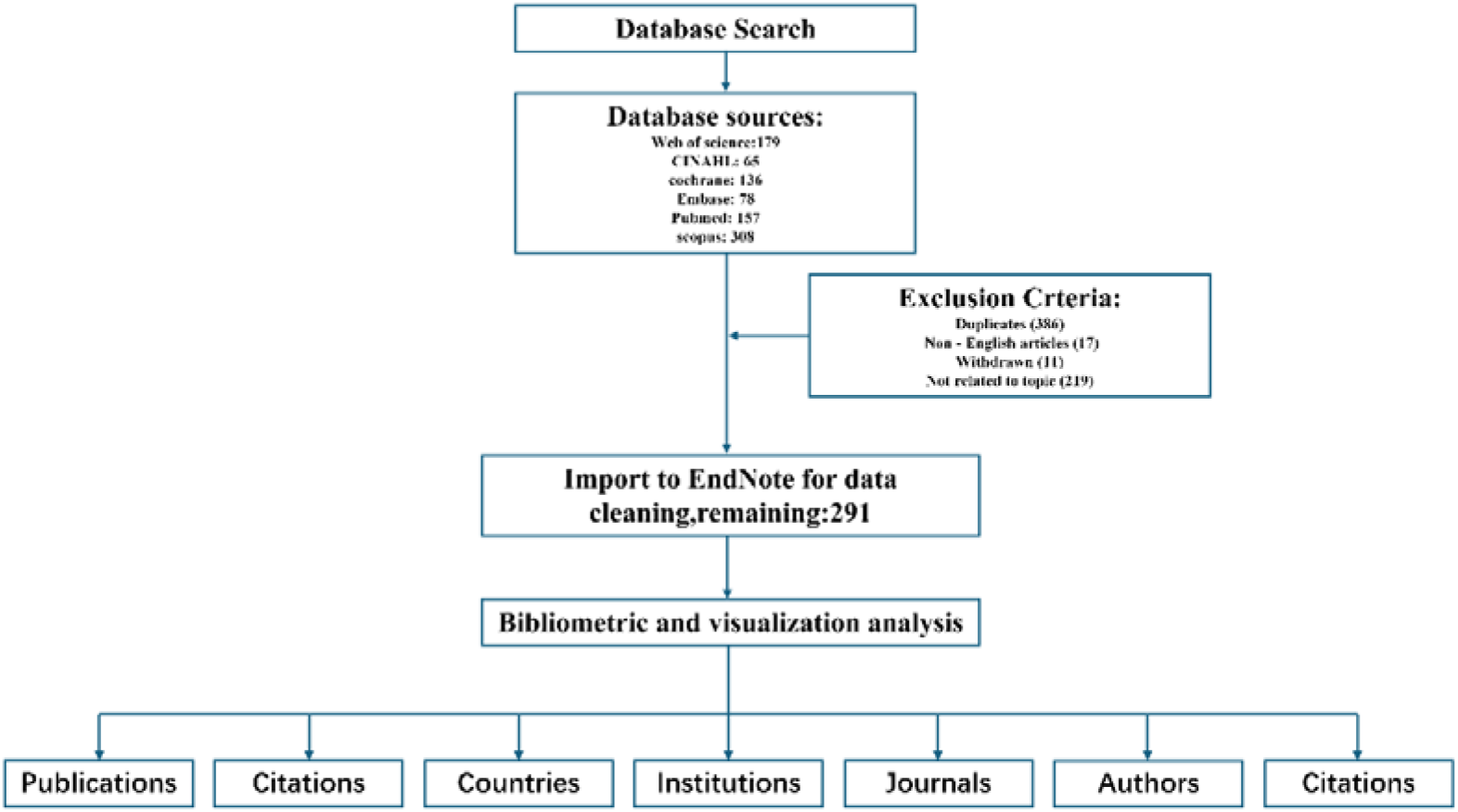
Publications screening flowchart.

### 2.2 Inclusion and exclusion criteria

Inclusion criteria: Articles or reviews published in peer-reviewed journals and indexed in at least one of the following databases: Web of Science Core Collection (WoSCC), Scopus, PubMed, Embase, Cochrane Library, and CINAHL Complete.

Exclusion criteria: (1) Non-English publications, (2) proceeding papers, early access articles, meeting abstracts, editorial materials, letters, book chapters, corrections, news items, reprints, or retracted publications, and (3) duplicate literature across databases.

### 2.3 Validation of Data

To validate the data, title searches were performed to confirm the absence of false positives, with assistance from two colleagues in the medical fields. The retrieved articles were cross-referenced with highly cited publications in Google Scholar to enhance the comprehensiveness of the analysis. The consistency of the metadata and alignment with active journals and prolific authors further confirmed the credibility of the search approach[22].

### 2.4 Study Selection and Bias

Information such as the title, authors, institutions, document type, journal, DOI, abstract, publication year, organization, citations, keywords, open-access status, and funding details were extracted and saved as a CSV file for subsequent analysis. Articles were screened to exclude those out of scope, and any missing information was completed. Duplicate records were removed based on the title and DOI using EndNote. To minimize potential bias, articles were ranked by citations, and comprehensive relevance screening was conducted to ensure accuracy and mitigate selection bias.

### 2.5 Scientific Literature Bibliometric Indicators

The cleaned dataset was exported to Microsoft Excel 365 and analyzed using R software (v.4.3.0) with the “bibliometrix” package. Advanced bibliometric indicators were calculated, including the total number of publications (TP), total citations (TC), average citations (AC), number of contributing authors (NCA), annual collaboration index (ACI), number of cited publications (NCP), citations per cited publication (CCP), collaboration index (CI), collaboration coefficient (CC), number of active years of publication (NAY), productivity per active year of publication (PAY), average citation per year (AC/Y), and author indices (h-index, g-index, and m-index). These indicators were measured and compared by publication year and citation patterns.

### 2.6 Software for bibliometric analysis

Bibliometric analysis employs mathematical and statistical methods to analyze literature database data, including publications, authors, institutions, and citations, generating knowledge maps to understand research landscapes and trends. Literature management was conducted using EndNote 21 for merging datasets, screening publications, and removing duplicates. Statistical analyses were performed using R software (version 4.3.0) with Bibliometrix, iGraph, and ggplot2 packages for bibliometric analysis, network visualization, and temporal trend analysis[23, 24]. Advanced visualizations were conducted using VOSviewer (1.6.20) and CiteSpace (5.7.R5). VOSviewer analyzed co-authorship networks among countries, institutions, journals, and authors, as well as keyword co-occurrence patterns[25]. CiteSpace performed burst detection analysis of keywords and literature co-citation analysis[26]. Thematic analysis employed bibliometric algorithms to categorize research themes into clusters based on conceptual similarity. Temporal evolution analysis tracked theme development across 2015-2025, calculating annual frequencies to identify research patterns.

Parameters were optimized for each approach: VOSviewer used full counting method with maximum 25 countries per article; CiteSpace employed time span 2015.01–2025.04 with annual slices, g-index25 (k = 25), LRF = 3.0, LBY = 8, e = 2.0; thematic analysis used adjusted keyword co-occurrence thresholds and semantic similarity algorithms for cluster identification.

## 3. RESULTS

### 3.1 Temporal Distribution Analysis

The temporal distribution of publications on operating room automation research demonstrates notable trends in research activity. As illustrated in **Figure 2**, publication output displays a fluctuating pattern with an overall upward trajectory from 2015 to 2024. The number of publications increased from 16 in 2015 to a peak of 49 in 2024, with 13 publications already recorded in the first four months of 2025 (**Table 1**). Particularly significant is the substantial growth observed during 2022-2024, when 132 articles were published (39 + 44 + 49), representing 45.4% of the total literature corpus of 291 publications. The research field experienced remarkable growth spurts, particularly in 2018 (100% growth rate), 2022 (69.57% growth rate), and steady increases in 2023-2024 (12.82% and 11.36% respectively). The early 2025 data (January-April) shows 13 publications, indicating sustained research momentum in the field.

**Figure 2.**
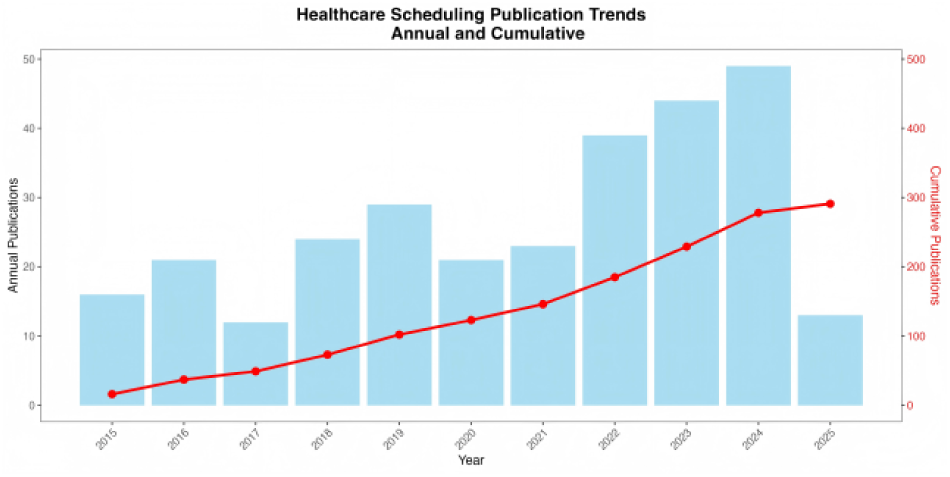
Temporal Distribution of Annual and Cumulative Publications in Operating Room Automation Research (2015-2025)

**Table 1.** Annual Publication Trends in Operating Room Automation Research (2015-2025)

| Year | Number of Publications | Cumulative Publications | Growth Rate (%) |
| --- | --- | --- | --- |
| 2015 | 16 | 16 | — |
| 2016 | 21 | 37 | 31.25 |
| 2017 | 12 | 49 | -42.86 |
| 2018 | 24 | 73 | 100.00 |
| 2019 | 29 | 102 | 20.83 |
| 2020 | 21 | 123 | -27.59 |
| 2021 | 23 | 146 | 9.52 |
| 2022 | 39 | 185 | 69.57 |
| 2023 | 44 | 229 | 12.82 |
| 2024 | 49 | 278 | 11.36 |
| 2025 | 13 | 291 | -73.47 |

### 3.2 Journal Distribution and Academic Impact Analysis

A total of 291 articles related to smart operating room research were published across 210 journals. Bradford’s Law analysis revealed a typical core-periphery structure: the core zone contained 29 journals publishing 98 articles (33.7%), Zone 2 included 85 journals with 97 articles (33.3%), and Zone 3 comprised 96 journals with 96 articles (33.0%) (**Figure 3B**, 3C). The International Journal of Computer Assisted Radiology and Surgery had the highest publication output (9 articles), followed by the Journal of Robotic Surgery (8 articles) and Lecture Notes in Computer Science (6 articles) (**Table 2**, **Figure 3A**). Regarding academic impact, Human Factors showed the highest average citations per article (27.67), followed by Anesthesia and Analgesia (4.38) and IJCARS (4.0), with an overall field average of 3.67 citations per article (**Figure 3D**). Journal co-citation network analysis revealed that 29 core journals formed four major academic clusters with 295 linkages and a total link strength of 320.75, primarily comprising clinical medicine journals (red cluster), computer informatics journals (green cluster), and intelligent technology journals (blue cluster), with the Journal of Robotic Surgery and IJCARS serving as central nodes that bridge interdisciplinary connections (**Figure 3E**).

**Figure 3.**
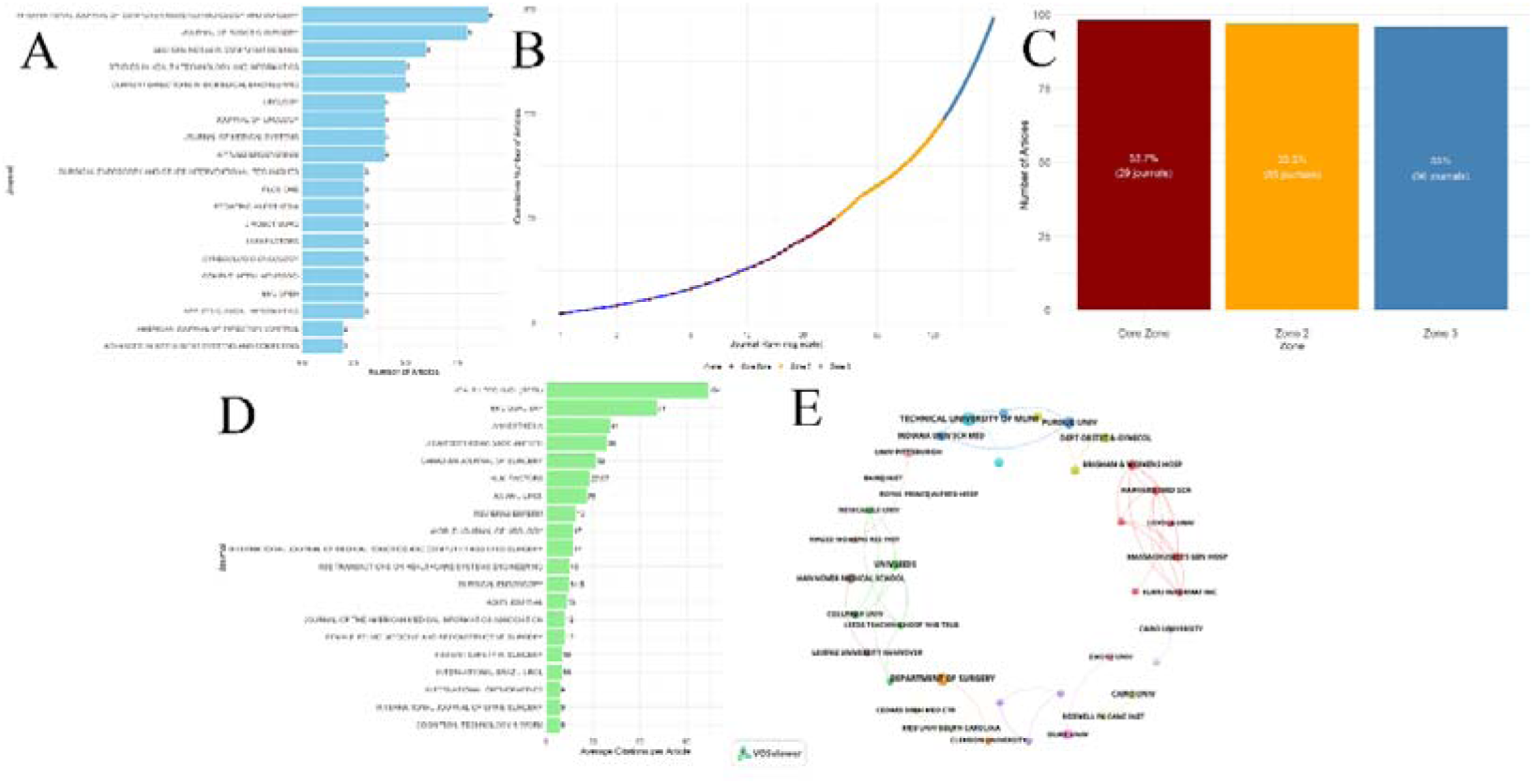
Journal Distribution and Impact Analysis in Operating Room Intelligence Research. (A) Top 20 journals ranked by publication output showing leading venues for research dissemination. (B) Bradford’s Law distribution demonstrating the characteristic exponential growth pattern of journal productivity. (C) Bradford’s zones distribution illustrating the core-periphery structure with three distinct zones. (D) Journal impact analysis displaying average citations per article across major publications. (E) Journal co-citation network visualization revealing interdisciplinary clustering patterns and collaborative relationships between different academic domains. [see Additional file 1]

**Table 2.**
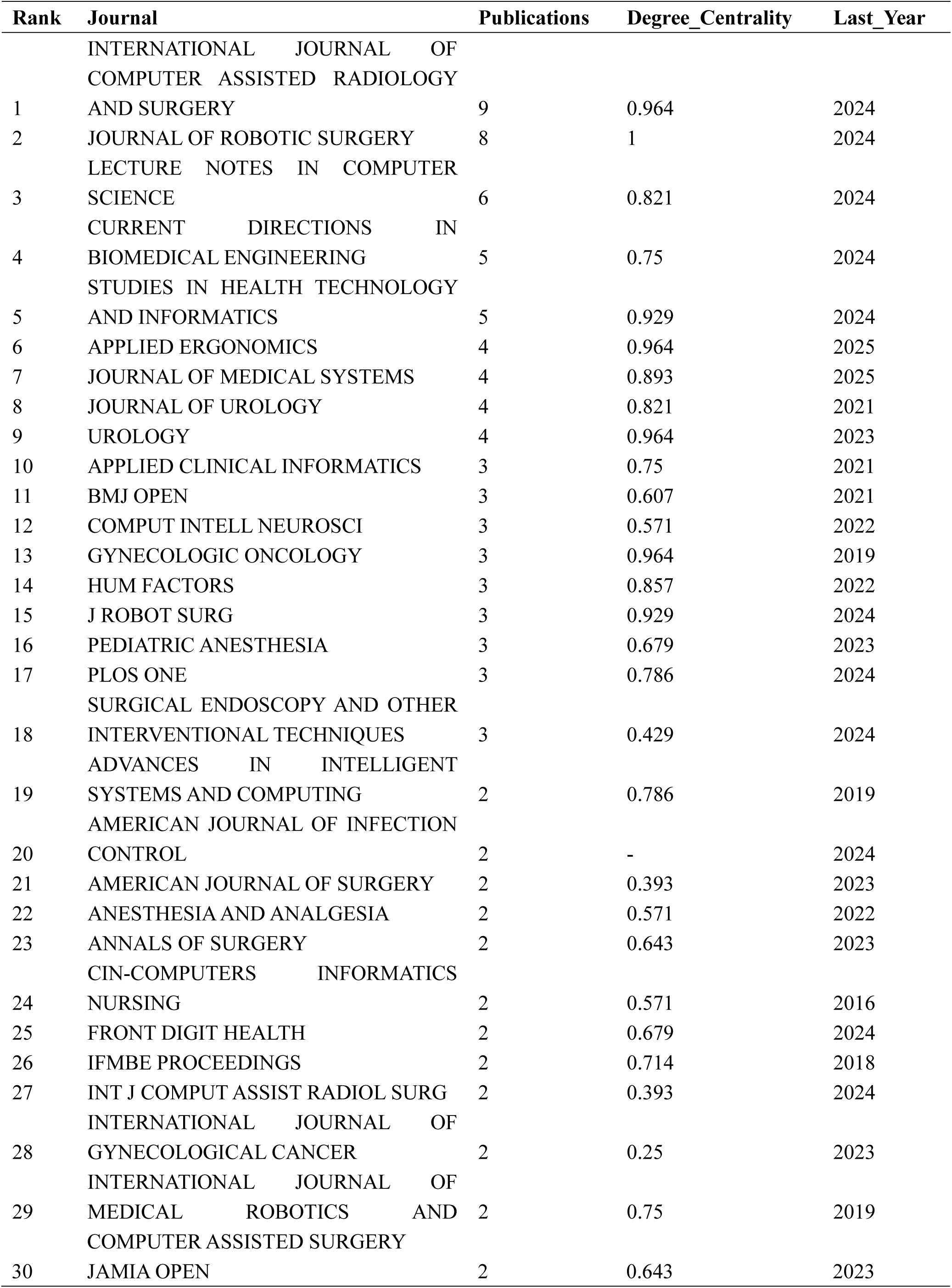
Comprehensive Analysis of Top Journals (2015-2025)

### 3.3 Geographical Distribution and International Collaboration Patterns

According to the literature analysis, the field of operating room intelligence demonstrates distinct geographical distribution characteristics and international collaboration patterns (Table 3, Figure 4). The United States leads with 109 publications, followed by the United Kingdom (33), Germany (33), and China (28) (Table 2, Figure 4A). VOSviewer-based international collaboration network analysis reveals a cooperative network encompassing 25 countries/regions with 7 clusters and a total link strength of 32 (Figure 4B), where the United States occupies the core position (weight: 109, links: 14) as the most important international collaboration hub, while China serves as a major Asian participant (weight: 28, links: 4) primarily collaborating with the US, UK, Japan, and other countries. The global publication distribution heat map (Figure 4C) further identifies three major research hotspots centered on North America (US), Europe (UK and Germany), and East Asia (China and Japan), with the network exhibiting a “core-periphery” structure characterized by geographical clustering, reflecting a regionalized collaboration model dominated by developed countries with gradually increasing participation from emerging economies.

**Figure 4.**
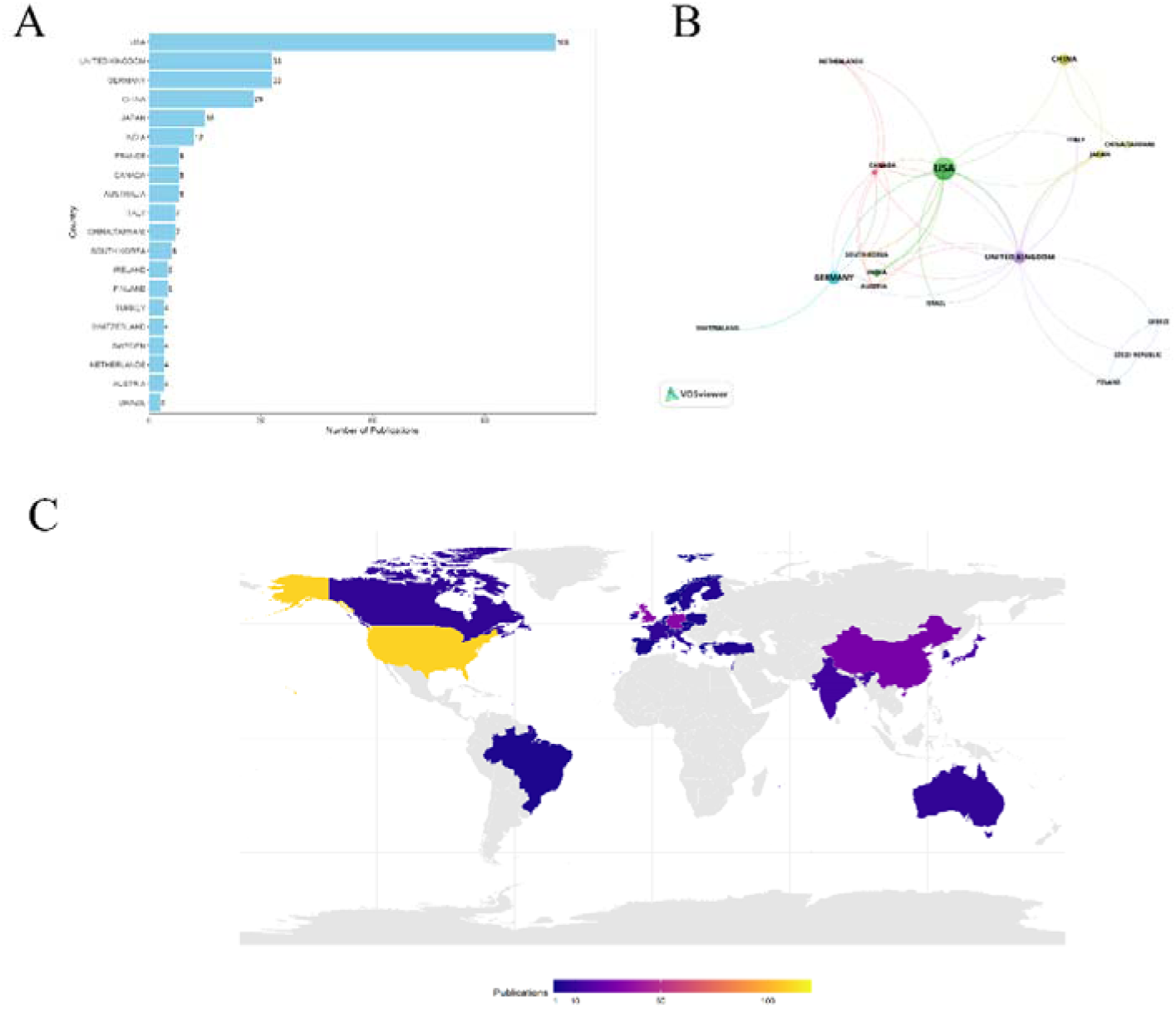
Global Distribution and International Collaboration Patterns in Operating Room Intelligence Research. (A) Top 20 countries ranked by publication output. (B) International collaboration network visualization showing cooperative relationships between countries. (C) Global publication distribution heat map illustrating geographical research intensity and identifying three major research clusters: North America, Europe, and East Asia. [see Additional file 1]

**Table 3.** Comprehensive Country Analysis of Operating Room Intelligence Research (2015-2025)

| Rank | Country | Publications | Degree Centrality | Betweenness Centrality |
| --- | --- | --- | --- | --- |
| 1 | USA | 109 | 26 | 23.85 |
| 2 | GERMANY | 33 | 8 | 24.24 |
| 3 | UNITED KINGDOM | 33 | 19 | 57.58 |
| 4 | CHINA | 28 | 5 | 17.86 |
| 5 | JAPAN | 15 | 7 | 46.67 |
| 6 | INDIA | 12 | 7 | 58.33 |
| 7 | AUSTRALIA | 8 | 2 | 25 |
| 8 | CANADA | 8 | 4 | 50 |
| 9 | FRANCE | 8 | 4 | 50 |
| 10 | CHINA(TAIWAN) | 7 | 3 | 42.86 |
| 11 | ITALY | 7 | 2 | 28.57 |
| 12 | SOUTH KOREA | 6 | 4 | 66.67 |
| 13 | FINLAND | 5 | 1 | 20 |
| 14 | IRELAND | 5 | 0 | 0 |
| 15 | AUSTRIA | 4 | 2 | 50 |
| 16 | NETHERLANDS | 4 | 3 | 75 |
| 17 | SWEDEN | 4 | 2 | 50 |
| 18 | SWITZERLAND | 4 | 3 | 75 |
| 19 | TURKEY | 4 | 1 | 25 |
| 20 | BRAZIL | 3 | 1 | 33.33 |
| 21 | ISRAEL | 3 | 3 | 100 |
| 22 | POLAND | 3 | 1 | 33.33 |
| 23 | SINGAPORE | 3 | 2 | 66.67 |
| 24 | BELGIUM | 2 | 1 | 50 |
| 25 | DENMARK | 2 | 0 | 0 |
| 26 | SPAIN | 2 | 0 | 0 |
| 27 | CZECH REPUBLIC | 1 | 1 | 100 |
| 28 | GREECE | 1 | 1 | 100 |
| 29 | NORWAY | 1 | 0 | 0 |

### 3.4 Institutional Distribution and Collaboration Patterns

Institutional analysis revealed that smart operating room research is predominantly concentrated in prestigious universities and medical institutions in Europe and North America (**Table 4**, **Figure 5A**). Technical University of Munich had the highest publication output (7 articles), followed by Department of Surgery (6 articles), Purdue University (5 articles), and Duke University (4 articles). Among the top 20 productive institutions, most published 3 articles, reflecting the relatively dispersed nature of research in this field. From a temporal perspective, Department of Surgery demonstrated the longest research span (2016-2025, 5 active years), while Technical University of Munich, despite having the highest publication count, showed a shorter research period (2022-2024, 3 active years), indicating the emergence of new research hotspots in the field. Institutional collaboration network analysis revealed that 39 major institutions formed 14 collaborative clusters with 39 linkages and a total link strength of 58, primarily comprising clusters of US East Coast medical schools (red cluster), European technical universities (blue cluster), and clinical medical institutions (green cluster), with Technical University of Munich and Department of Surgery serving as key nodes connecting research forces across different regions and disciplines (**Figure 5B**).

**Figure 5.**
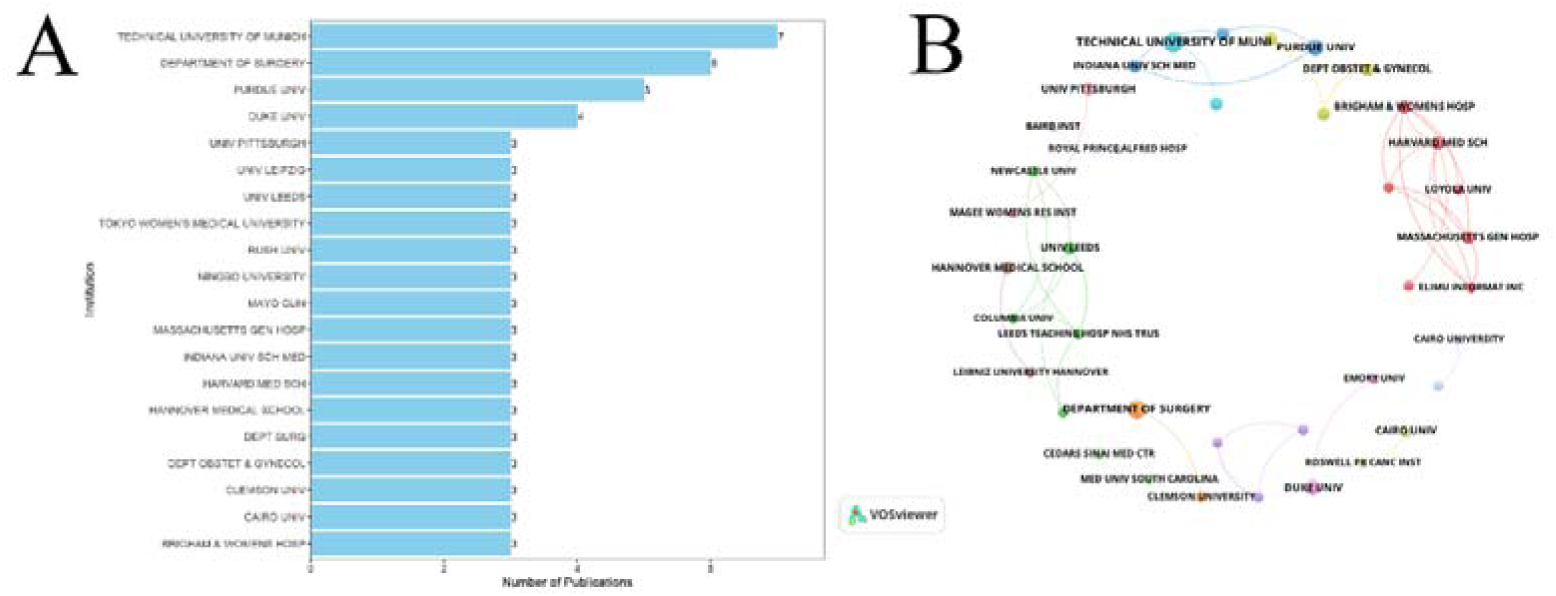
Institutional Distribution and Collaboration Patterns in Operating Room Intelligence Research. (A) Top 20 most productive institutions ranked by publication output, demonstrating the concentration of research in leading universities and medical centers. (B) Institutional collaboration network visualization showing inter-institutional partnerships and collaborative relationships across different geographical regions and academic disciplines. [see Additional file 1]

**Table 4.** Comprehensive Analysis of Top Institutions (2015-2025)

| Rank | Short_Name | Publication<br>s | Degree_Centrality | Active_Years |
| --- | --- | --- | --- | --- |
| 1 | TECHNICAL UNIVERSITY OF MUNICH | 7 | 0.026 | 3 |
| 2 | DEPARTMENT OF SURGERY | 6 | 0.026 | 5 |
| 3 | PURDUE UNIV | 5 | 0.053 | 4 |
| 4 | DUKE UNIV | 4 | 0.026 | 3 |
| 5 | BRIGHAM & WOMENS HOSP | 3 | 0.132 | 3 |
| 6 | CAIRO UNIV | 3 | 0.026 | 3 |
| 7 | CLEMSON UNIV | 3 | 0.079 | 1 |
| 8 | DEPT OBSTET & GYNECOL | 3 | 0.053 | 3 |
| 9 | DEPT SURG | 3 | 0.079 | 3 |
| 10 | HANNOVER MEDICAL SCHOOL | 3 | 0.026 | 3 |
| 11 | HARVARD MED SCH | 3 | 0.132 | 3 |
| 12 | INDIANA UNIV SCH MED | 3 | 0.053 | 2 |
| 13 | MASSACHUSETTS GEN HOSP | 3 | 0.105 | 3 |
| 14 | MAYO CLIN | 3 | 0.053 | 3 |
| 15 | NINGBO UNIVERSITY | 3 | - | 1 |
| 16 | RUSH UNIV | 3 | - | 3 |
| 17 | TOKYO WOMEN'S MEDICAL UNIVERSITY | 3 | - | 3 |
| 18 | UNIV LEEDS | 3 | 0.105 | 3 |
| 19 | UNIV LEIPZIG | 3 |  | 3 |
| 20 | UNIV PITTSBURGH | 3 | 0.026 | 2 |
| 21 | UNIVERSITY HOSPITAL RECHTS DER ISAR | 3 | 0.026 | 3 |
| 22 | VANDERBILT UNIV | 3 | - | 3 |
| 23 | HOSPITAL UNIVERSITY | 2 | - | 1 |
| 24 | BAIRD INST | 2 | 0.026 | 2 |
| 25 | CAIRO UNIVERSITY | 2 | 0.026 | 2 |
| 26 | CEDARS SINAI MED CTR | 2 | 0.026 | 2 |
| 27 | CHAN SCHOOL OF MEDICINE | 2 | 0.053 | 2 |
| 28 | CHINA MED UNIV | 2 | - | 1 |
| 29 | CLEMSON UNIVERSITY | 2 | 0.026 | 2 |
| 30 | COLUMBIA UNIV | 2 | 0.079 | 1 |

### 3.5 Author Productivity and Collaboration Network Analysis

The distribution of author publications in the field of operating room intelligence exhibits typical long-tail characteristics, conforming to Lotka’s Law in bibliometrics. According to author publication statistics (**Table 5**), WILHELM D leads with 7 publications, establishing himself as the most influential scholar in this field; BERNHARD L follows closely with 6 publications; WAGNER L and XIANG W tie for third place with 5 publications each. Additionally (**Figure 6A**). Lotka’s Law validation results show an author productivity distribution slope of −3.915, indicating that a small number of highly productive authors contribute a large volume of literature while most authors publish only 1-2 articles (**Figure 6B**). The author collaboration network analysis constructed using VOSviewer software reveals a network comprising 6 main clusters with a total link strength of 32, where WILHELM D serves as the network’s core node with 14 connections, making him the most important collaboration hub in this field (**Figure 6C**). The collaboration network exhibits distinct clustering characteristics, with the blue cluster centered on WILHELM D including scholars such as BERNHARD L, WAGNER L, KOLB S, and JELL A, forming the most tightly connected research team. Other clusters are represented in different colors, including the red cluster (ABRAHAM J, MIGUCHI M, etc.) and the green cluster (BEN ABDALLAH A, KRONZER A, etc.), reflecting the diversified research team organizational patterns and relatively stable academic collaboration relationships in this field.

**Figure 6.**
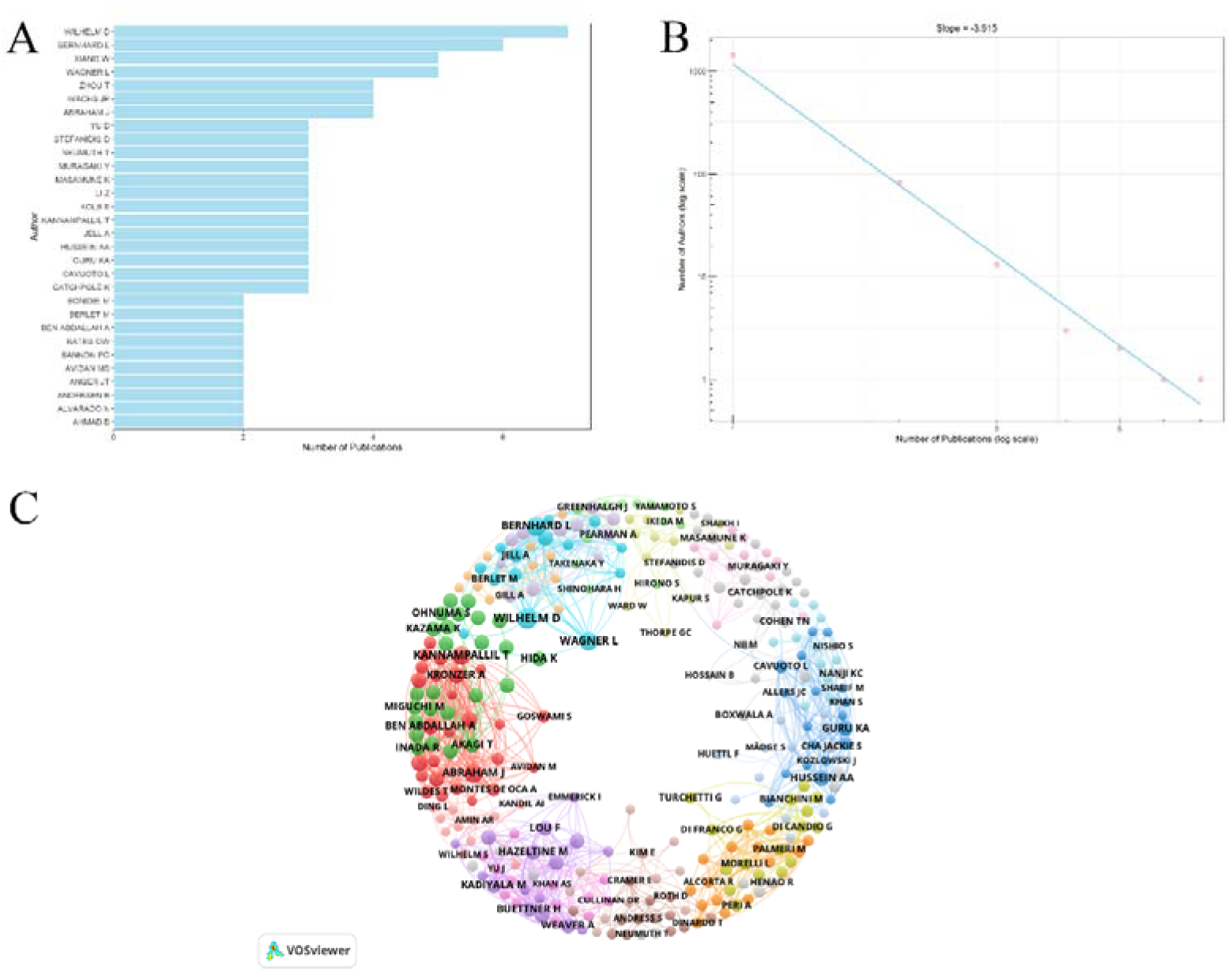
Author Productivity Analysis and Collaboration Network in Operating Room Intelligence Research. (A) Productive authors in healthcare scheduling ranked by publication output. (B) Author productivity distribution following Lotka’s Law with slope = −3.915, demonstrating the characteristic long-tail distribution. (C) Author collaboration network visualization showing research clusters and collaborative relationships between scholars. [see Additional file 1]

**Table 5.**
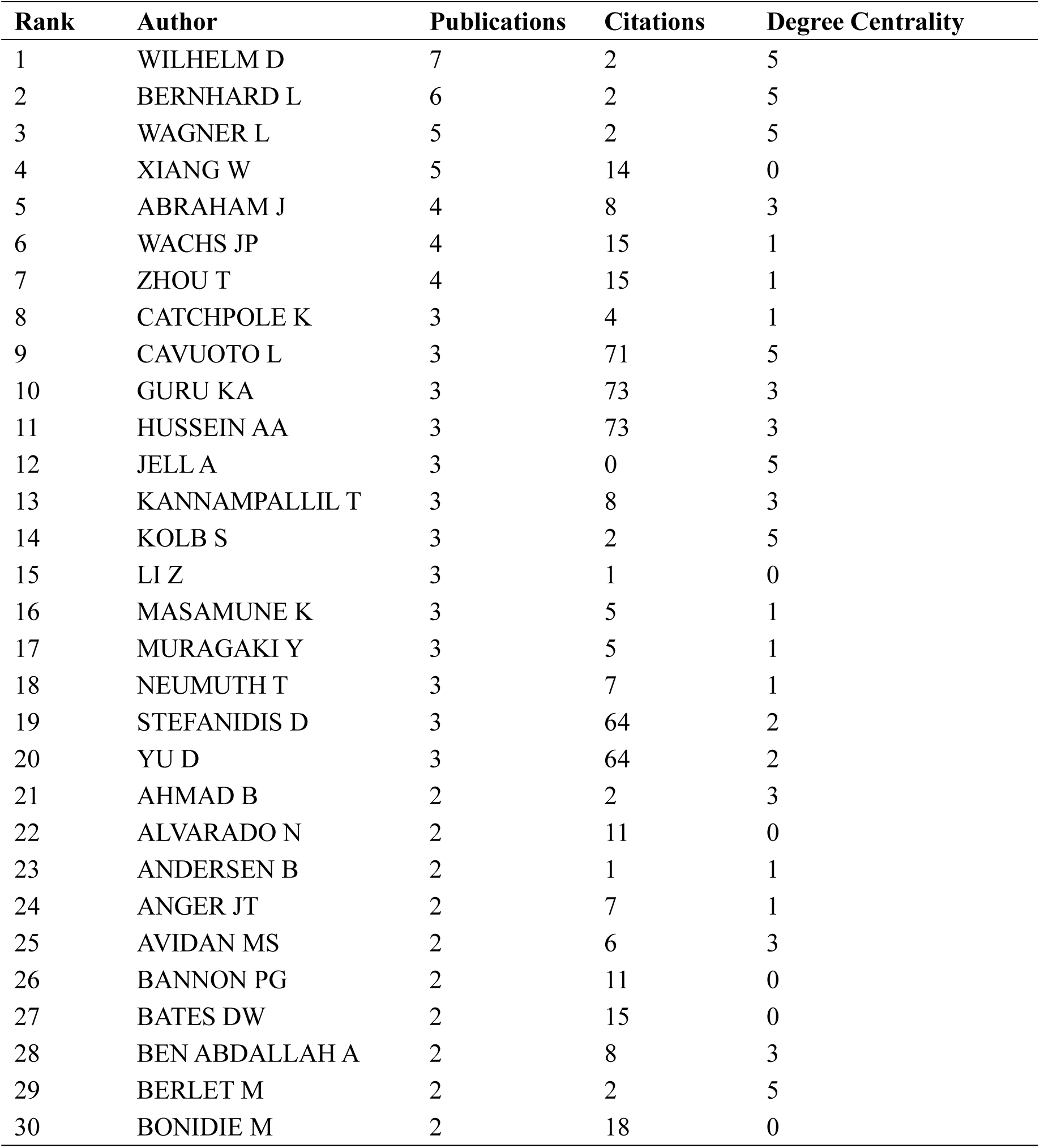
Productivity and Network Centrality (2015-2025)

| Rank | Author | Publications | Citations | Degree Centrality |
| --- | --- | --- | --- | --- |
| 1 | WILHELM D | 7 | 2 | 5 |
| 2 | BERNHARD L | 6 | 2 | 5 |
| 3 | WAGNER L | 5 | 2 | 5 |
| 4 | XIANG W | 5 | 14 | 0 |
| 5 | ABRAHAM J | 4 | 8 | 3 |
| 6 | WACHS JP | 4 | 15 | 1 |
| 7 | ZHOU T | 4 | 15 | 1 |
| 8 | CATCHPOLE K | 3 | 4 | 1 |
| 9 | CAVUOTO L | 3 | 71 | 5 |
| 10 | GURU KA | 3 | 73 | 3 |
| 11 | HUSSEIN AA | 3 | 73 | 3 |
| 12 | JELL A | 3 | 0 | 5 |
| 13 | KANNAMPALLIL T | 3 | 8 | 3 |
| 14 | KOLB S | 3 | 2 | 5 |
| 15 | LI Z | 3 | 1 | 0 |
| 16 | MASAMUNE K | 3 | 5 | 1 |
| 17 | MURAGAKI Y | 3 | 5 | 1 |
| 18 | NEUMUTH T | 3 | 7 | 1 |
| 19 | STEFANIDIS D | 3 | 64 | 2 |
| 20 | YU D | 3 | 64 | 2 |
| 21 | AHMAD B | 2 | 2 | 3 |
| 22 | ALVARADO N | 2 | 11 | 0 |
| 23 | ANDERSEN B | 2 | 1 | 1 |
| 24 | ANGER JT | 2 | 7 | 1 |
| 25 | AVIDAN MS | 2 | 6 | 3 |
| 26 | BANNON PG | 2 | 11 | 0 |
| 27 | BATES DW | 2 | 15 | 0 |
| 28 | BEN ABDALLAH A | 2 | 8 | 3 |
| 29 | BERLET M | 2 | 2 | 5 |
| 30 | BONIDIE M | 2 | 18 | 0 |

### 3.6 Keyword Analysis and Research Hotspot Evolution

Keyword analysis revealed core themes and developmental trends in smart operating room research (**Table 6**, **Figure 7**). Among 100 high-frequency keywords, “OPERATING ROOM” had the highest occurrence (100 times), followed by “ROBOTIC SURGERY” (89 times), “NURSING” (67 times), “TEAMWORK & COMMUNICATION” (44 times), and “ARTIFICIAL INTELLIGENCE” (40 times), reflecting the field’s emphasis on both technological applications and humanistic care (**Figure 7A**). Regarding network centrality, “ROBOTIC SURGERY” demonstrated the highest degree centrality (0.863) and strength centrality (0.756), with “NURSING” (degree centrality 0.897, strength centrality 0.801) following closely, indicating these topics occupy central positions in the knowledge network. Keyword co-occurrence network analysis showed that 50 core keywords formed six major research clusters with 585 linkages and a total link strength of 1467, primarily focusing on operating room management, robotic technology, nursing practice, patient safety, quality improvement, and intelligent algorithms (**Figure 7B**). Citation burst analysis identified 23 keywords with burst characteristics, with early hotspots including “operating room scheduling” (2017), and “multi-objective optimization” (2017), while recent emerging trends encompass “deep learning” (2020-2023), “natural language processing” (2023), and “robotic scrub nurse” (2023-2025) (**Figure 7C**, 7D).

**Figure 7.**
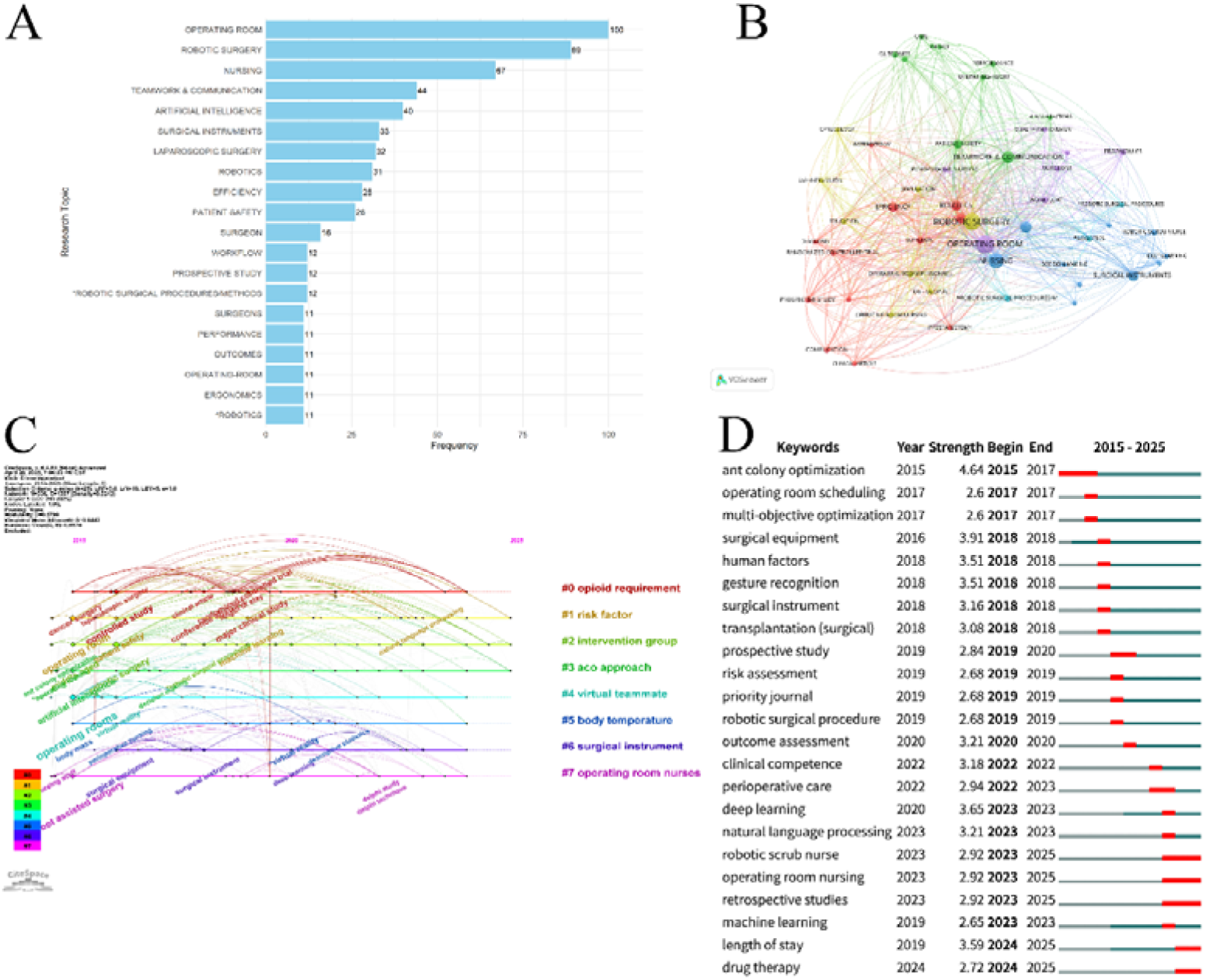
Keyword Analysis and Research Hotspot Evolution in Operating Room Intelligence Research. (A) Top 20 research topics in healthcare scheduling ranked by frequency, showing the most prominent research themes. (B) Keyword co-occurrence network visualization revealing six major research clusters and their interconnections. (C) Keyword evolution timeline demonstrating the temporal development of research themes from 2015-2025. (D) Top 23 keywords with the strongest citation bursts, highlighting emerging trends and research hotspots over time. [see Additional file 1] Table 6. Comprehensive Analysis of Top Keywords (2015-2025)

**Table 6.**
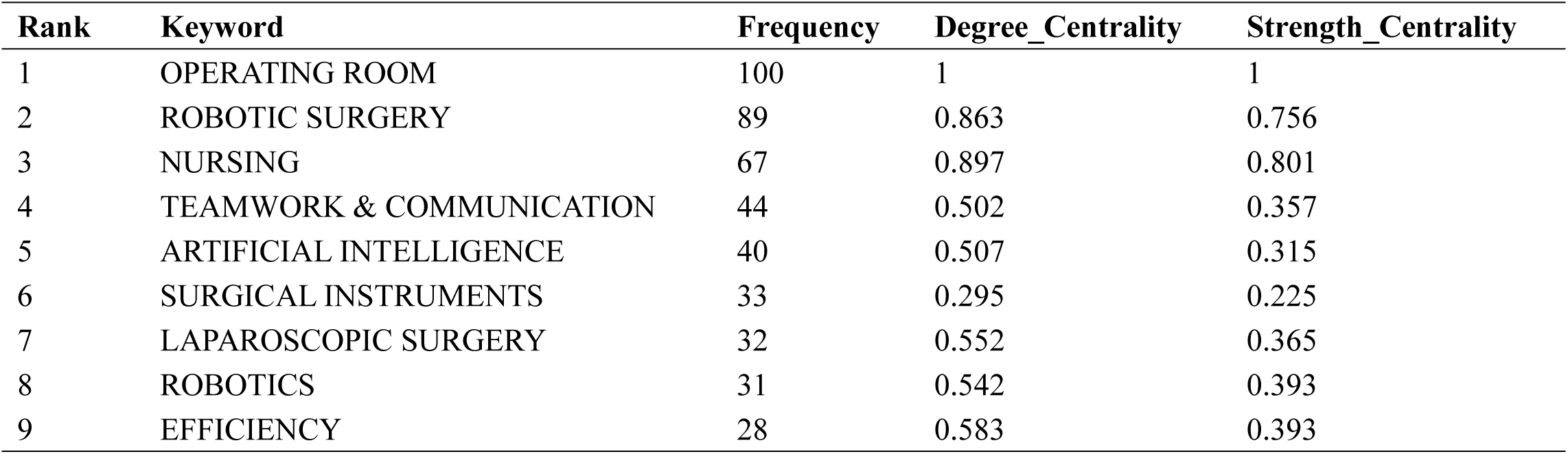
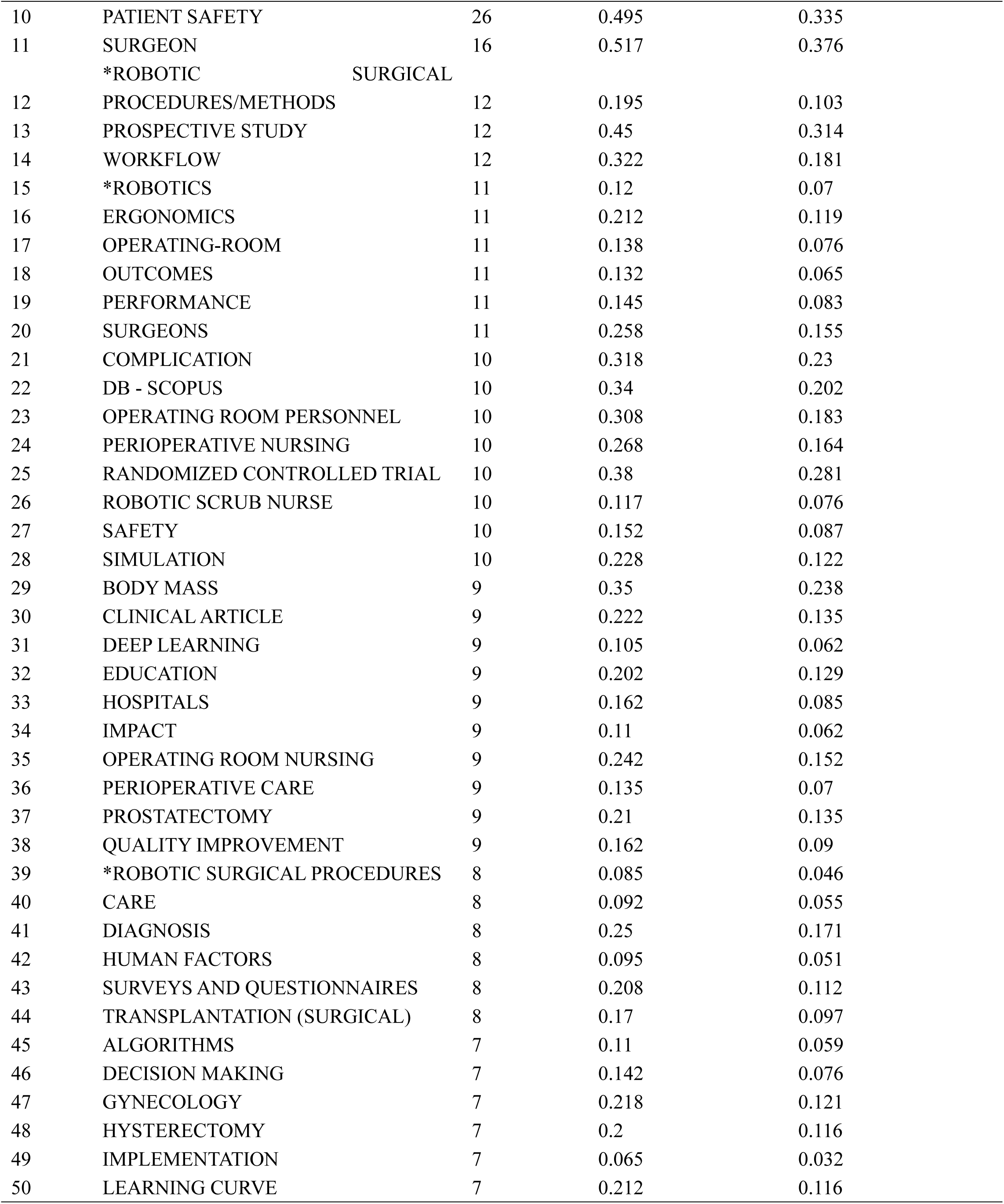
Comprehensive Analysis of Top Keywords (2015-2025)

### 3.7 Thematic Evolution and Research Development Trends

Thematic evolution analysis identified six core research clusters in the smart operating room field: Surgical Robotics & Equipment, Nursing Staff & Scheduling, Surgical Settings & Workflow, Safety Quality & Risk Management, Intelligent Algorithms & Decision Support, Communication & Collaboration, and Information Systems & Data Management (**Figure 8A**). The thematic relationship network revealed that Surgical Robotics & Equipment represents the largest cluster with the strongest inter-thematic connections, while Communication & Collaboration and Intelligent Algorithms & Decision Support developed relatively independently. Temporal evolution trends showed dynamic characteristics across 2015-2025: Surgical Robotics & Equipment peaked in 2019 (33 occurrences), Surgical Settings & Workflow was most active in 2019 (28 occurrences), Nursing Staff & Scheduling experienced significant growth in 2019 (25 occurrences), while Intelligent Algorithms & Decision Support started strong in 2015 (10 occurrences) before stabilizing (**Figure 8B**). Heat map analysis further revealed that Surgical Robotics & Equipment continuously intensified during 2016-2019, Nursing Staff & Scheduling experienced concentrated emergence in 2018-2019, and Safety Quality & Risk Management has gained increasing importance in recent years, reflecting the field’s evolution from technology-oriented towards human-machine collaboration and integrated quality-safety considerations (**Figure 8C**, **8D**).

**Figure 8.**
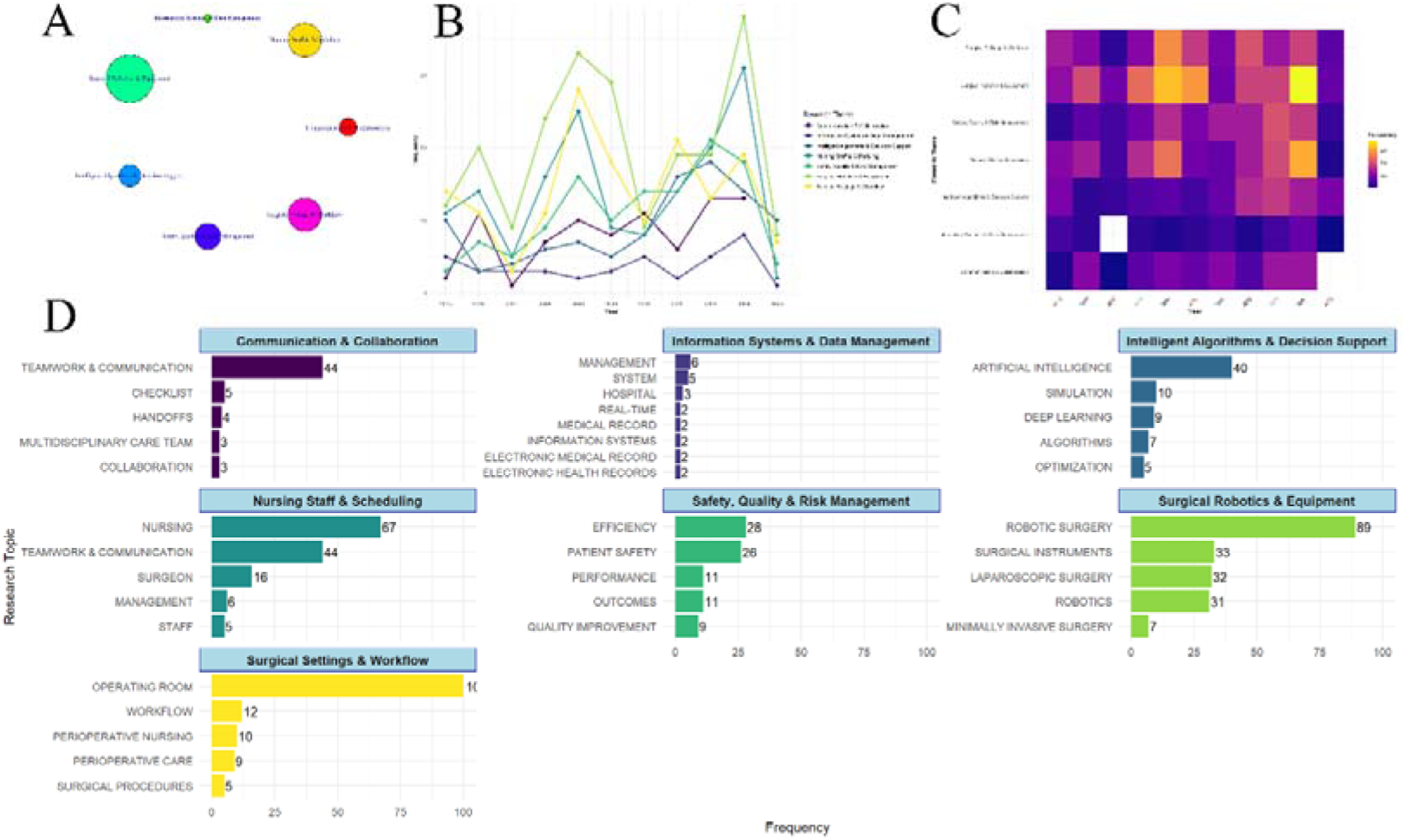
Thematic Evolution and Research Development Trends in Operating Room Intelligence Research. (A) Relationships between research themes showing stronger connections only, illustrating the interconnectedness of major research clusters. (B) Evolution of research themes over time (2015-2025) demonstrating the temporal dynamics and intensity changes of different research areas. (C) Research themes in intelligent operating room nursing categorized by different domains including communication & collaboration, information systems & data management, nursing staff & scheduling, safety quality & risk management, surgical robotics & equipment, and surgical settings & workflow. (D) Evolution of research themes in intelligent OR nursing presented as a heat map, revealing the temporal concentration and development patterns of various research themes. [see Additional file 1]

## 4. Discussion

By examining relevant literature, we conducted a comprehensive bibliometric analysis of smart OR nursing studies[27, 28]. The bibliometric patterns observed in these studies provide insights into the opportunities and barriers facing the integration of smart technologies into OR nursing[29], suggesting key implementation challenges and identifying impediments that may prevent the translation of research findings into clinical practice.

### 4.1 Temporal Distribution and Journal Publication Patterns

Publications grew from 16 (2015) to 49 (2024), with 2022-2024 representing 45.4% of total output (132 out of 291 publications). The field demonstrated significant growth momentum, particularly with notable increases in 2018 (100% growth rate), 2022 (69.57% growth rate), and sustained growth in 2023-2024 (12.82% and 11.36% respectively). Concurrently, 291 articles scattered across 210 journals following Bradford’s Law, with International Journal of Computer Assisted Radiology and Surgery leading (9 publications) and Human Factors achieving highest impact (27.67 citations per article) against a field average of 3.67. The co-citation network reveals four interdisciplinary clusters bridged by Journal of Robotic Surgery and IJCARS. This temporal-journal evolution demonstrates a coherent pattern of field maturation from concentrated foundational work to diversified specialization. The inverse relationship between growing publication volume and declining citations directly corresponds to the wide journal scatter, where researchers transition from citing core foundational texts to drawing from increasingly diverse and specialized literature across multiple disciplinary venues[30]. The early research period (2015-2017) established foundational knowledge in operating room automation, while the publication surge in 2022-2024 reflects the field’s maturation and practical implementation across diverse healthcare settings. The journal distribution pattern reinforces this evolutionary trajectory. Bradford’s Law compliance with relatively even zone distribution indicates that operating room intelligence has not yet consolidated around dominant publication venues, reflecting the field’s expansion across traditional disciplinary boundaries[31]. Human Factors’[32] exceptional citation performance (27.67) demonstrates that foundational human-centered design principles maintain broad relevance even as research diversifies, while the modest field average (3.67) suggests that specialized technical applications receive more limited cross-field recognition.

The four-cluster co-citation structure (clinical medicine, computer informatics, intelligent technology) mirrors the temporal shift toward specialized applications, where researchers naturally gravitate toward domain-specific publication venues. While Journal of Robotic Surgery[33] [34]and IJCARS[35, 36] serve as crucial interdisciplinary bridges, this fragmentation across both time and publication landscape creates knowledge silos that challenge integrated field development. The temporal concentration of recent publications combined with journal dispersion suggests the field requires new mechanisms for knowledge synthesis to maintain coherent advancement despite increasing specialization.

### 4.2 Geographic Distribution and Institutional Concentration Patterns

The dominance of developed countries—particularly the US (109 publications), UK (33), and Germany (33)—directly corresponds to institutional concentration among prestigious technical universities and medical centers, with Technical University of Munich (7 publications), Purdue University (5), and Duke University (4) leading research output. Meanwhile, emerging economies like China, despite substantial research capacity (28 publications), maintain more limited international connections (4 links) and correspondingly fewer leading institutions in global networks.This geographical-institutional convergence reflects critical infrastructure requirements that extend beyond research capacity alone. Leading countries benefit from established regulatory pathways like the FDA’s (Food and Drug Administration) medical device approval framework and tight integration between elite academic institutions, world-class medical centers, and advanced technology companies[37]. The institutional collaboration network, organized around US East Coast medical schools, European technical universities, and clinical medical institutions, mirrors the geographical clustering pattern where regional proximity facilitates complex interdisciplinary partnerships essential for this field[38, 39]. The temporal evolution reveals how institutional and geographical advantages reinforce each other. While established centers like Department of Surgery demonstrate sustained engagement (2016-2025), new hotspots such as Technical University of Munich’s intense recent activity (2022-2024) reflect expanding but still concentrated research infrastructure. This pattern suggests that specialized resources—advanced robotics laboratories, clinical testing environments, regulatory expertise, and substantial capital investment—remain concentrated in elite institutions within developed countries, creating barriers for emerging economies to achieve full integration into global research networks[40, 41].

However, even these resource-rich geographical regions and institutions face substantial implementation barriers. The relatively dispersed publication pattern among institutions (most contributing only 3 articles) indicates that even well-funded centers struggle to maintain sustained research programs due to high capital costs, complex clinical workflows, and surgical team resistance to automation[42, 43]. This suggests that successful operating room intelligence development requires not just technological innovation but fundamental healthcare system transformation—a challenge that transcends individual institutional or national capabilities.

### 4.3 Author Productivity and Collaboration Network Patterns

The author productivity analysis reveals a highly concentrated landscape with WILHELM D leading at 7 publications and serving as the central collaboration hub (14 connections), followed by a small core group including BERNHARD L, WAGNER L, and XIANG W. The Lotka’s Law validation (slope = −3.915) confirms extreme concentration where few authors drive knowledge creation while most contribute only 1-2 articles. The collaboration network shows 6 distinct clusters with WILHELM D functioning as the primary knowledge broker. This steep concentration reflects the field’s specialized nature, requiring substantial expertise and resources that few researchers can sustain consistently. The pattern indicates operating room intelligence research remains in an early consolidation phase, where pioneer researchers establish foundational knowledge and collaborative frameworks. WILHELM D’s central position enables cross-cluster knowledge transfer but creates potential bottlenecks for field advancement[44–46]. The distinct clustering patterns suggest research teams organize around complementary expertise rather than geographical proximity, forming stable collaborative relationships essential for complex interdisciplinary projects. However, this heavy dependence on individual researchers poses both advantages—enabling deep specialization and sustained collaboration—and risks for field development if key contributors become unavailable.

### 4.4 Keyword Analysis and Thematic Evolution Patterns

Among 100 high-frequency keywords, “OPERATING ROOM” leads with 100 occurrences, followed by “ROBOTIC SURGERY” (89), “NURSING” (67), “TEAMWORK & COMMUNICATION” (44), and “ARTIFICIAL INTELLIGENCE” (40). Regarding network centrality, “NURSING” demonstrates the highest degree centrality (0.897) and strength centrality (0.801), with “ROBOTIC SURGERY” following closely (degree centrality 0.863, strength centrality 0.756), indicating these topics occupy central positions in the research landscape.

Keyword co-occurrence network analysis revealed that 50 core keywords formed six major research clusters with 585 linkages and a total link strength of 1467, primarily focusing on operating room management, robotic technology, nursing practice, patient safety, quality improvement, and intelligent algorithms. Citation burst analysis identified 23 keywords with burst characteristics, revealing temporal evolution from early focus on “operating room scheduling” (2017) and “multi-objective optimization” (2017) to recent emphasis on “deep learning” (2020-2023), “natural language processing” (2023), and “robotic scrub nurse” (2023-2025).

This evolution demonstrates the field’s progression through distinct phases: from operating room scheduling and optimization approaches to AI integration, and from technology-centered approaches to human-machine collaboration systems. The high centrality of both “ROBOTIC SURGERY” and “NURSING” reflects the field’s recognition that successful automation requires deep clinical workflow integration rather than purely technological solutions[47, 48]. The citation burst progression from scheduling optimization to sophisticated AI applications demonstrates accelerating technological adoption, while the emphasis on teamwork, communication, and quality improvement indicates field maturation toward comprehensive human-machine collaborative systems[49].

### 4.5 Future Research Directions and Policy Implications

Future research must prioritize comparative effectiveness studies evaluating intelligent technologies’ impact on patient outcomes and operational efficiency across diverse healthcare settings. Cross-national validation models should assess technology feasibility while accounting for infrastructure and resource variations. Implementation science methodologies must examine organizational factors affecting adoption, including culture, leadership, and workflow redesign, alongside health equity assessments to prevent exacerbating healthcare disparities[50, 51].Global regulatory harmonization is critical for broader implementation. Current frameworks inadequately address AI systems that continuously learn and evolve, requiring new approaches balancing innovation with patient safety. Evidence standards must encompass dynamic AI performance beyond traditional device metrics, while professional scope guidelines should clarify roles in AI-augmented environments and liability frameworks address legal concerns impeding adoption[52, 53]. Meanwhile, enhancing collaboration between researchers, clinicians, policymakers, and technology developers is essential. International partnerships should explore culturally appropriate strategies for resource-limited countries, ensuring global healthcare equity. Interdisciplinary collaborations must integrate technical, economic, and organizational expertise, while industry-academia partnerships should develop scalable implementation models adaptable across diverse healthcare contexts.

### 4.6 Study Limitations

This analysis has several limitations affecting implementation planning implications. Academic literature may exhibit publication bias toward positive results, potentially overrepresenting successful implementations while underreporting failed pilots. The focus on English-language publications from developed countries may miss important implementation experiences from diverse healthcare contexts.

Additionally, temporal lag in academic publishing means current implementation challenges may not be reflected in peer-reviewed literature, potentially limiting findings’ relevance for immediate decision-making.

## 5. Conclusions

This study explores operating room intelligence research through comprehensive bibliometric analysis, providing an overview of its evolution, identifying emerging trends, and evaluating its progression from 2015-2025.

Key research hotspots include robotic surgery, nursing automation, AI integration, and human-machine collaboration systems. The field has evolved from operational optimization (2017) to deep learning applications (2020-2023) and robotic scrub nurse development (2023-2025). Future research should focus on comparative effectiveness studies, cross-national validation models, and implementation science methodologies addressing organizational factors and health equity concerns.

International collaboration reveals significant concentration, with the United States leading (109 publications) alongside the UK, Germany, and China. However, emerging economies maintain limited international connections, highlighting global research inequities. Enhanced collaboration between researchers, clinicians, policymakers, and technology developers is essential for translating technological advancements into practical clinical applications. The field faces substantial implementation barriers despite technological progress. High capital costs, complex clinical workflows, and resistance to automation challenge even well-resourced institutions. Moving forward, addressing regulatory harmonization for AI systems, developing evidence standards for dynamic AI performance, and establishing professional scope guidelines will be crucial for broader adoption.

Future research should prioritize global health equity, ensuring operating room intelligence benefits diverse healthcare contexts rather than exacerbating existing disparities. Interdisciplinary partnerships must integrate technical, economic, and organizational expertise while developing scalable implementation models. The field requires enhanced knowledge synthesis mechanisms to prevent disciplinary fragmentation and maintain coherent advancement toward comprehensive human-machine collaborative systems that improve patient outcomes and healthcare delivery worldwide.

## Data Availability

Data presented in this study are included in the article/Supplementary Materials; further inquiries can be directed to the corresponding author.

## Author information

## Author notes

Yu Yang and Zhili Rao share first authorship.

## Contributions

Yu Yang was responsible for research conception, data collection, data analysis, and writing the main manuscript text; Zhili Rao was responsible for data collection, data analysis, and reviewing of the manuscript; Wen Zheng was responsible for methodology guidance and reviewing of the manuscript; Yaling Wang was responsible for project supervision, scientific research design, and reviewing of the manuscript. All other authors contributed to data collection, clinical practice, and reviewing of the manuscript. All authors read and approved the final manuscript.

## Ethics declarations

## Ethics approval and consent to participate

Not applicable.

## Consent for publication

All authors have approved the submission. This manuscript has not been submitted or published elsewhere.

## Abbreviations

The following abbreviations are used in this manuscript:

IJCARS: INTERNATIONAL JOURNAL OF COMPUTER ASSISTED RADIOLOGY AND SURGERY
OR: Operating Room
FDA: Food and Drug Administration
AI: artificial intelligence

